# Severe, unrecognized sleep disturbances in patients with cirrhosis diagnosed with a portable electroencephalogram device

**DOI:** 10.1101/2025.03.28.25324860

**Authors:** Atsushi Uchiyama, Hiroteru Kamimura, Suguru Miida, Hiroki Maruyama, Takafumi Tonouchi, Jaehoon Seol, Toshio Kokubo, Tomohiro Okura, Yusuke Watanabe, Naruhiro Kimura, Hiroyuki Abe, Akira Sakamaki, Takeshi Yokoo, Shuji Terai

## Abstract

**Goals:** We aimed to examine patients with liver cirrhosis and compare their sleep characteristics with those of healthy controls using innovative new electroencephalogram models.

**Background:** Sleep disturbance is a common disorder in patients with cirrhosis. Many previous studies have been conducted on the topic using questionnaire methods. However, polysomnographic quantitative assessments remain challenging because they are complicated and difficult to implement at the clinical research level.

**Study:** We recruited patients with early liver disease with a low probability of causing sleep disturbances, body mass index (BMI) ≤31 kg/m^2^, history of alcohol use, itching, and no severe encephalopathy. Using propensity score matching among 20 patients with liver cirrhosis and 95 healthy older adults, the sleep structure of 18 patients with cirrhosis was analyzed and compared with that of 18 healthy adults matched for age, sex, and BMI. Sleep structure in both the groups was assessed using data obtained from a portable electroencephalogram (EEG) device, including total sleep time, sleep latency, wakefulness after sleep onset, sleep efficiency, sleep stages, and rapid eye movement (REM) latency. Sleep stages included REM and non-rapid eye movement (non-REM) stages (stages N1, N2, and N3). Self-report questionnaires were administered to the participants to confirm subjective and objective sleep statuses.

**Results:** Questionnaire responses revealed that neither group reported any severe sleep disturbances. Regarding sleep structure assessment using a portable EEG device, patients with cirrhosis exhibited longer sleep latency, suboptimal wakefulness after sleep onset, and poorer sleep efficiency than healthy controls. In patients with cirrhosis, the total time and percentage of N1 sleep were longer; total time and percentage of REM sleep were shorter; and REM latency was prolonged.

**Conclusions:** A high frequency of severely unrecognized sleep disturbances was observed in patients with cirrhosis using home portable polysomnography.

## Introduction

Cirrhosis has become increasingly prevalent globally, partly due to population growth and aging. Deaths due to cirrhosis accounted for 2.4% of global deaths in 2017, compared with 1.9% in 1990. Sleep-wake disturbance is common in patients with cirrhosis, with 50–80% of them experience disturbances five times higher than do healthy individuals. However, polysomnography, the gold standard for assessing sleep architecture, is extremely difficult to conduct in research due to instrumental complexity and insurance coverage; moreover, knowledge about it is rather limited [1].

Medical history and physical examinations can help identify patients at risk of cirrhosis. Patients with cirrhosis frequently experience muscle cramps (64%), pruritus (39%), poor sleep quality (63%), and sexual dysfunction (53%) [2]. However, many of the risk factors, such as diabetes or alcohol use, along with the symptoms, are neither sensitive nor specific [3].

This study evaluated comparison of pure cirrhotic patients with healthy controls patients based on the American Academy of Sleep Medicine’s (AASM) 2017 criteria.

## Materials and Methods

### Ethics statement

The study protocol conformed to the guidelines of the Declaration of Helsinki (as revised in Fortaleza, Brazil, October 2013) and was approved by the institutional Ethics Review Board of the Niigata University (approval number: 2021-0274). Additionally, we used the data from the study by Seol et al. (2022) [4] approved by the Ethics Committee of the University of Tsukuba (approval number: R02-211).

### Study design, participants, and settings

#### Inclusion criteria

Patients with cirrhosis were enrolled from Niigata University Graduate School of Medical and Dental Sciences, while healthy control individuals were enrolled between 2021 and 2023 from University of Tsukuba. We included outpatients who (1) had not used sleeping pills in the past 6 months and (2) had body mass index ≤31 kg/m^2^ (cutoff for sleep apnea diagnosis) [5,6]. Participation was voluntary; all participants provided written consent.

#### Exclusion criteria

We excluded patients (1) with a history of a major shunt, mainly spleno-renal (which might cause hyperammonemia), as observed on computed tomography [7]; (2) with moderate, severe, or very severe itching, defined based on the Verbal Rating Scale score (none, mild, moderate, severe, or very severe) and considered to interfere with daily life activities [8]; (3) with clinically apparent encephalopathy or psychiatric comorbidities; and (4) with experience of alcohol use disorder or reported daily alcohol use of 20 g within the last 6 months.

### Reference population

Overall, 95 healthy volunteers (75% male, mean age = 75.9 years, mean body mass index = 23.6 kg/m^2^) were recruited from the University of Tsukuba. The same tool was used for data collection. Using propensity score matching based on sex, age, and body mass index, the study participants were matched with a caliper of 1. No patient had reported evidence of alcohol abuse, chronic liver disease, or neurological/psychiatric disorders; consumed >20 g of alcohol daily; or took prescription drugs.

Patients with Child-Pugh class A cirrhosis who met the above criteria were fitted with a portable electroencephalogram (EEG) device. After explaining its operating procedure, a trial fitting was conducted to eliminate operational problems. Data were collected over three days. In patients with cirrhosis, serum ammonia, hepatic reserve, and Epworth Sleepiness Scale (ESS) tests were performed. Data were then compared with those of the reference group.

### Determining sleep stages in adults

The AASM published a scoring manual on non-REM sleep (Table 1), which is now recommended for determining sleep stages [9]. REM sleep is classified into shallow sleep (Stage N1 and Stage N2) and deep sleep (Stage N3).

**Table 1.** Determination of Sleep Stages in Adults.

| Determination of sleep stages in adults |  |  |
| --- | --- | --- |
| AASM scoring manual 2007 |  |  |
| Awakening |  | Stage W |
| Non-REM sleep | Shallow sleep | Stage N1 |
|  |  | Stage N2 |
|  | Deep sleep<br>slow wave sleep | Stage N3 |
| REM sleep |  | Stage R |

AASM, American Academy of Sleep Medicine; REM, rapid eye movement; non-REM
(NREM), non-rapid eye movement.
Stage W: Wakefulness
Stage N1: Light sleep (Stage 1 of NREM sleep)
Stage N2: Intermediate sleep (Stage 2 of NREM sleep)
Stage N3: Deep sleep or slow wave sleep (Stage 3 of NREM sleep)
Stage R: REM sleep

### Characteristics of the sleep stages

The characteristics of each sleep stage are summarized in Table 2. Sleep stages are determined in epochs of 30 seconds. Stage N1 is characterized by a lower amplitude and frequency of alpha activity (8–13 Hz) than awake EEG, with a predominance of 4–7 Hz. Stage N2 is characterized by the presence of sleep spindles and K-complexes, with less than 20% slow wave activity. Stage N3 has slow wave activity (0.5–2.0 Hz, amplitude >75 µV) for >20% of an epoch. Stage R includes low-frequency background activity similar to that of Stage N1, with a frequency of α activity (8–13 Hz) less than 50% of the time and a dominance of 4–7 Hz. Stage R is also similar to Stage N1 in showing low-frequency background activity, with sawtooth waves, REMs on electro-oculography, and reduced muscle tone compared with non-REM sleep on electromyography. The electromyogram is characterized by REMs and a decrease in muscle tone compared with non-REM sleep [10].

**Table 2.**
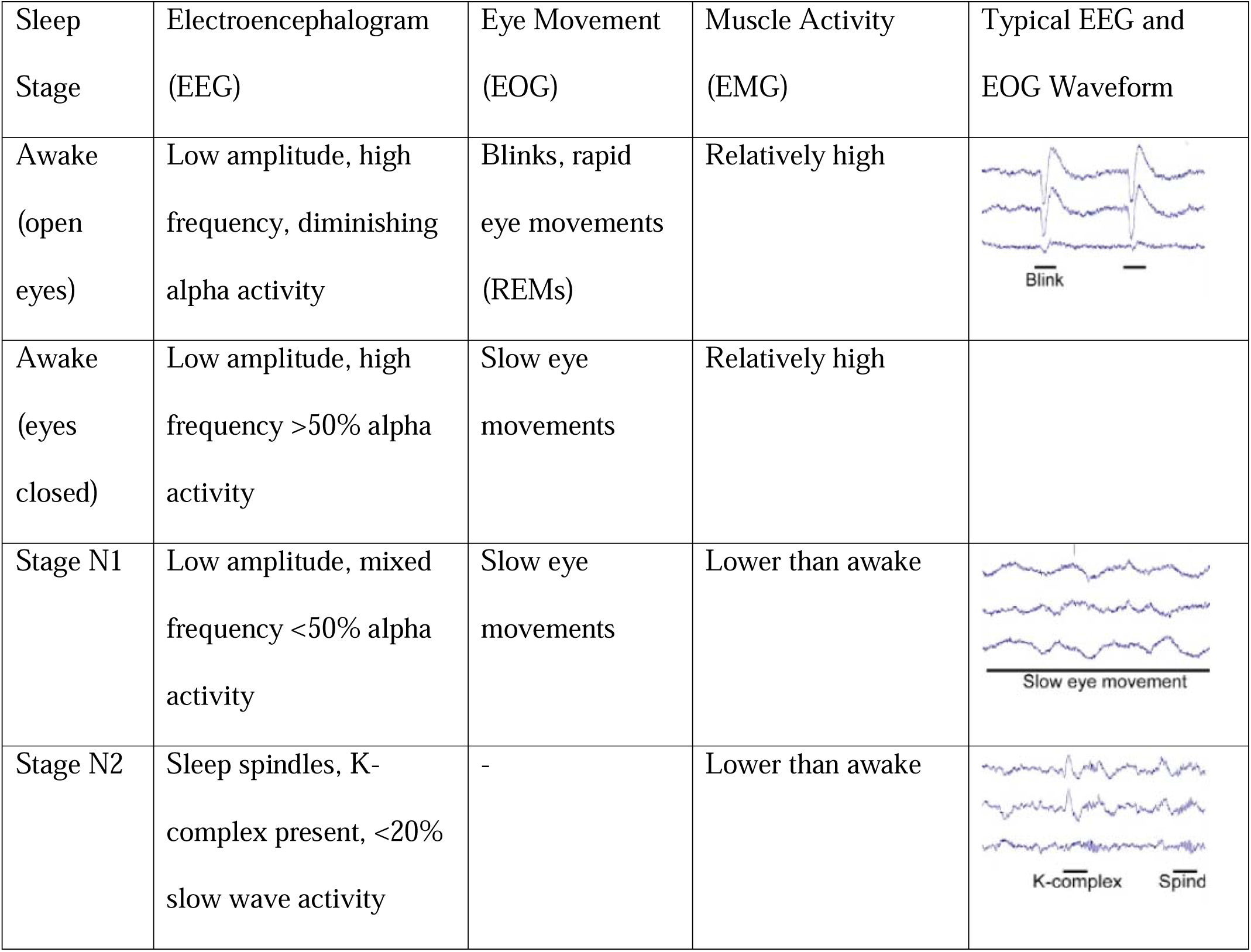
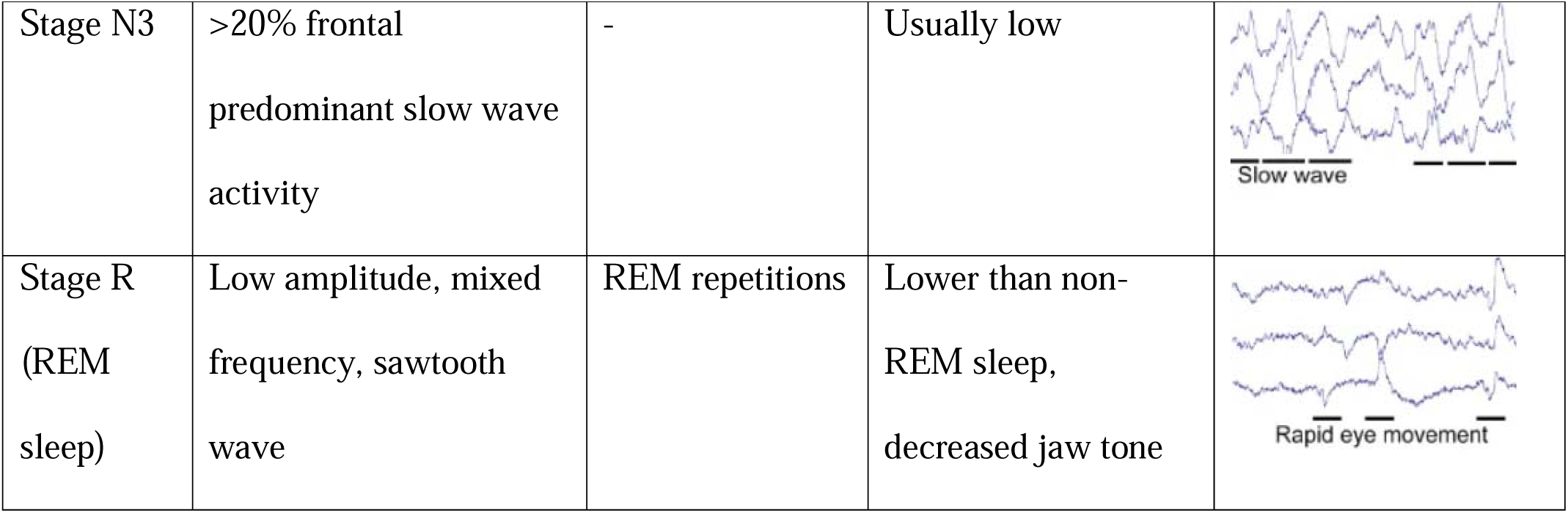
Characteristics of Sleep Stages.

| Sleep Stage | Electroencephalogram (EEG) | Eye Movement (EOG) | Muscle Activity (EMG) | Typical EEG and EOG Waveform |
| --- | --- | --- | --- | --- |
| Awake (open eyes) | Low amplitude, high frequency, diminishing alpha activity | Blinks, rapid eye movements (REMs) | Relatively high |  |
| Awake (eyes closed) | Low amplitude, high frequency >50% alpha activity | Slow eye movements | Relatively high |  |
| Stage N1 | Low amplitude, mixed frequency <50% alpha activity | Slow eye movements | Lower than awake |  |
| Stage N2 | Sleep spindles, K-complex present, <20% slow wave activity | - | Lower than awake |  |
| Stage N3 | >20% frontal<br>predominant slow wave<br>activity | - | Usually low |  |
| Stage R<br>(REM<br>sleep) | Low amplitude, mixed<br>frequency, sawtooth<br>wave | REM repetitions | Lower than non-REM sleep,<br>decreased jaw tone |  |

### ESS

Conventional sleepiness was assessed using the ESS (Fig 1). Participants were asked to rate their likelihood of “falling asleep” in eight different situations during the day on a scale of 0 (unlikely) to 3 (very likely); the eight scores were summed up to result in a total score between 0 and 24. A score higher than 11 points was considered as strong daytime sleepiness [11].

**Fig 1.**
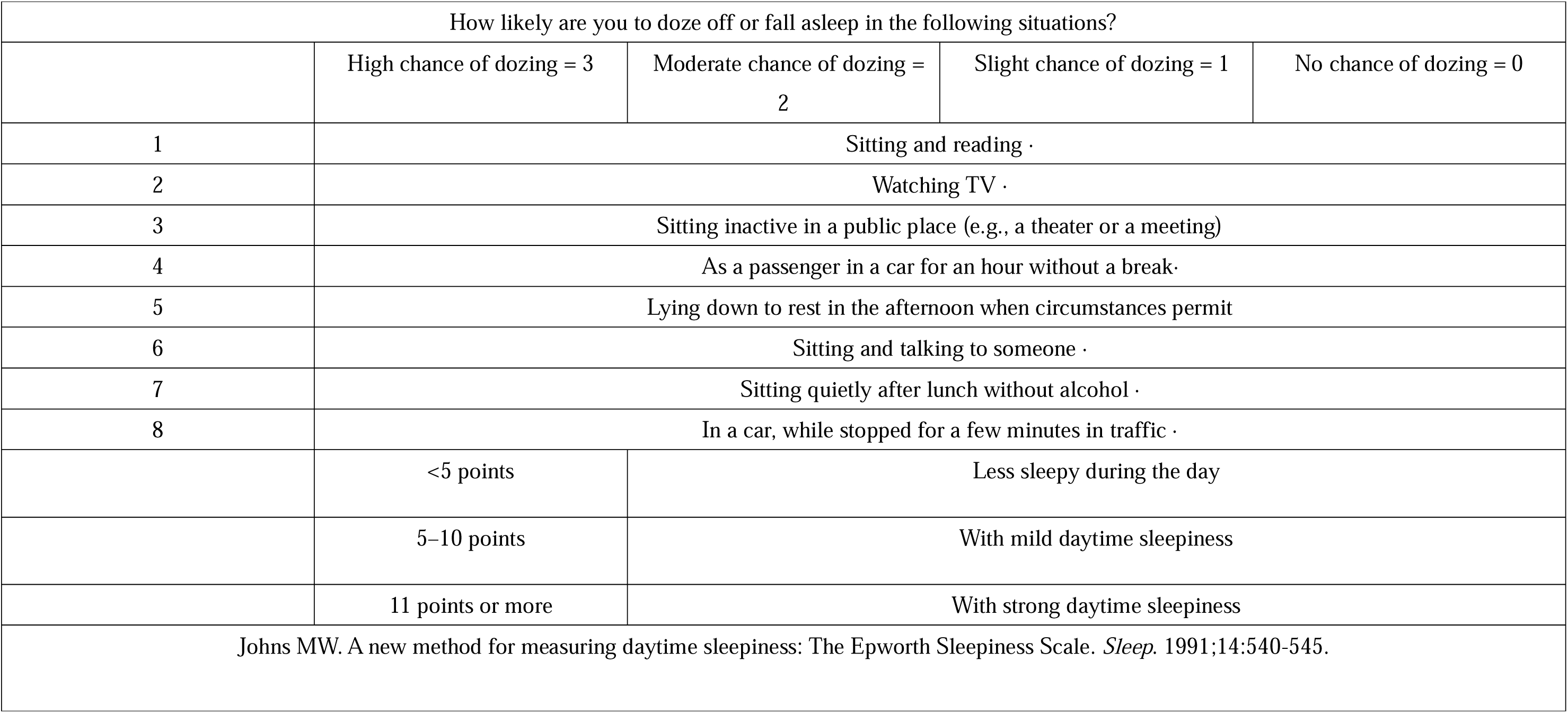
The Epworth Sleepiness Scale.

### Sleep variables

#### Portable EEG device and self-reported sleep parameters

Objective daily sleep parameters were recorded using a portable EEG device (Insomnograf; S’UIMIN Inc., Tokyo, Japan) (Fig 2). The device was light (162 g) and easy to attach and remove, various montages are possible, and EOG and EMG components can be displayed by changing the filtering conditions. Compared with a typical polysomnography device, the portable EEG device exhibited a high concordance rate of 0.80 in healthy controls and 0.78 in patients with sleep apnea [12].

**Fig 2.**
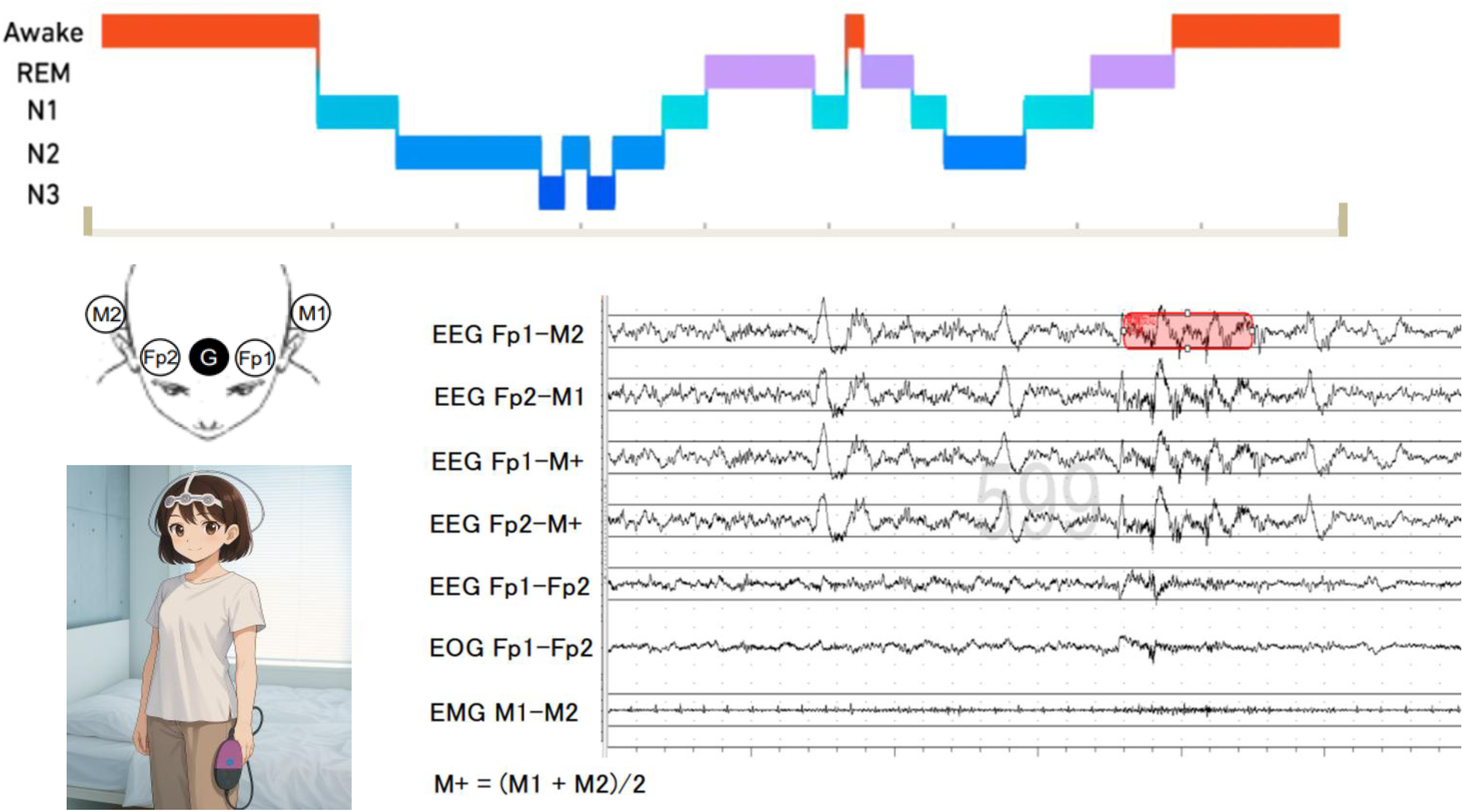
Features of the portable electroencephalogram device Potential data to be measured.

The participants were instructed to attach the adhesive electrodes to their heads after showering and push the recording button immediately before going to bed and after waking up. Bedtime and the last waking up time in the morning were determined using these data. Recording ended when the participants either pressed the power button or inserted a charging cable. We crosschecked the bedtimes and wake-up times from the portable EEG device with those of the self-reported questionnaires to ensure that data were collected even when the participants forgot to push the button after waking up. The recording system of this device comprised four EEG electrodes (Fp1, Fp2, M1, and M2) and one reference electrode (Fpz), positioned according to the 10–20 system. The montage was combined with four electroencephalogram derivations (Fp1–M2, Fp2–M1, Fp1–average M, and Fp2–average M). Here, Fp1–Fp2 and Fp2–Fp1 were used for left and right electro-oculography, respectively, and M1–M2 for chin electromyography to analyze sleep staging. The records were scored at 30-s intervals to classify sleep stages in the following categories: wakefulness (stage W); non-REM (stage N); N1, N2, and N3; and REM. Measurements during sleep onset latency were classified as stage W. Moreover, REM latency was measured until the first REM stage after sleep onset. Waking up after sleep onset was defined according to standard criteria (AASM 2017).

### Statistical analysis

Propensity score matching was performed based on a previous study, with the caliper set to 1.0 [13]. All statistical analyses were performed using analysis of variance, followed by paired-samples *t*-tests comparing the means of both matched groups. All analyses were performed using the IBM SPSS Statistics software (Version 29.0; IBM Corporation, Armonk, NY, USA) for Windows, and Microsoft Excel version 2408 was used to illustrate Fig 1. Statistical significance was set at *P* < 0.05 (two-tailed).

## Results

### Patient characteristics

Propensity score matching was used to select 18 from the 20 patients with cirrhosis and 18 from the 95 healthy controls. Overall, 18 participants were included (15 male individuals, mean age = 72.9 years). Furthermore, 12 participants were diagnosed with metabolic dysfunction-associated steatohepatitis; 8 were treated for liver cancer. All participants had a Child-Pugh A reserve capacity, and no patient had more than first-degree encephalopathy according to the West Haven criteria classification, with a mean NH3 value of 70.3 ± 5.5 µg/dL. The ESS was 6.2 ± 2.3 points; no patient showed severe sleep disturbance in the ESS results (Table 3).

**Table 3.**
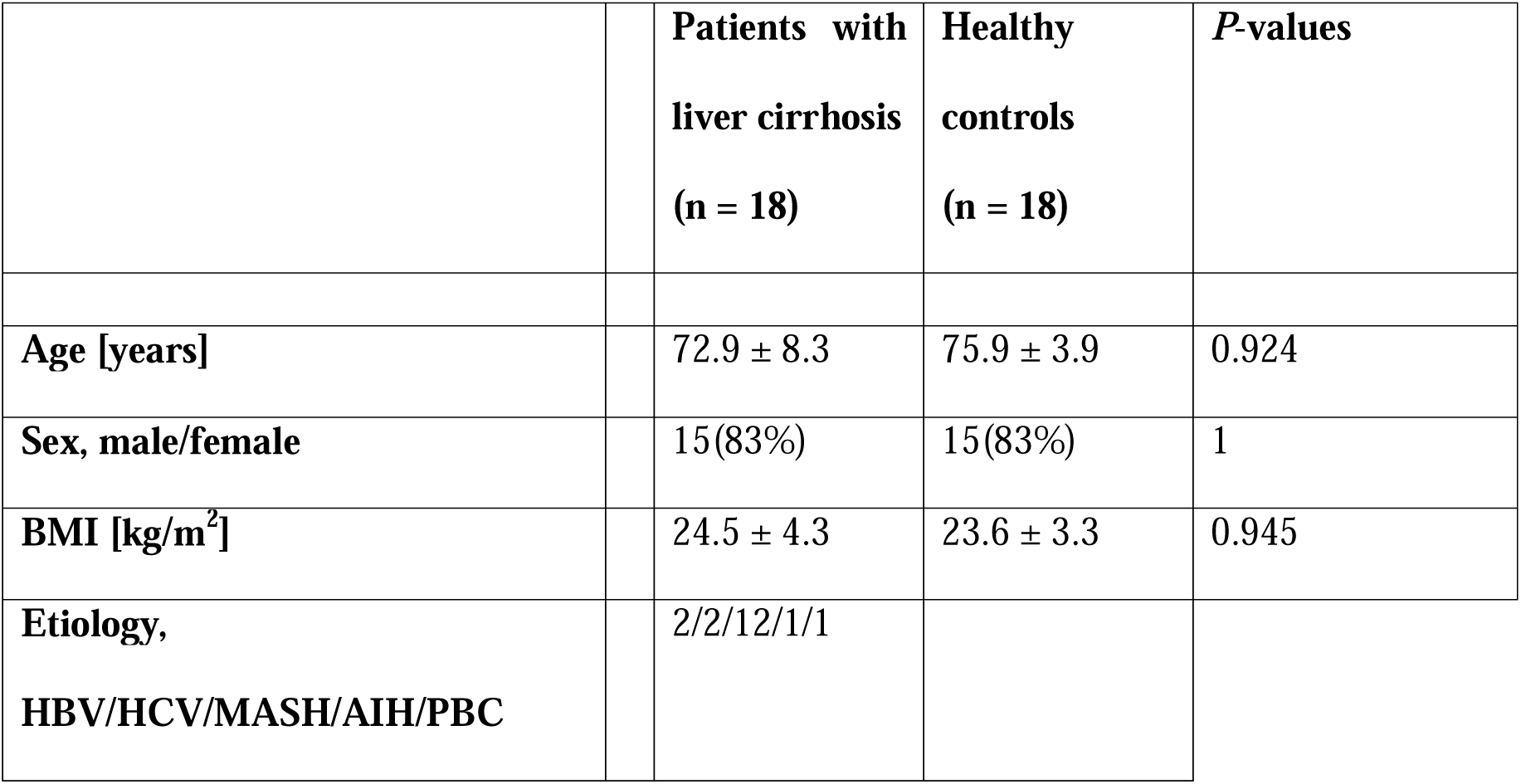
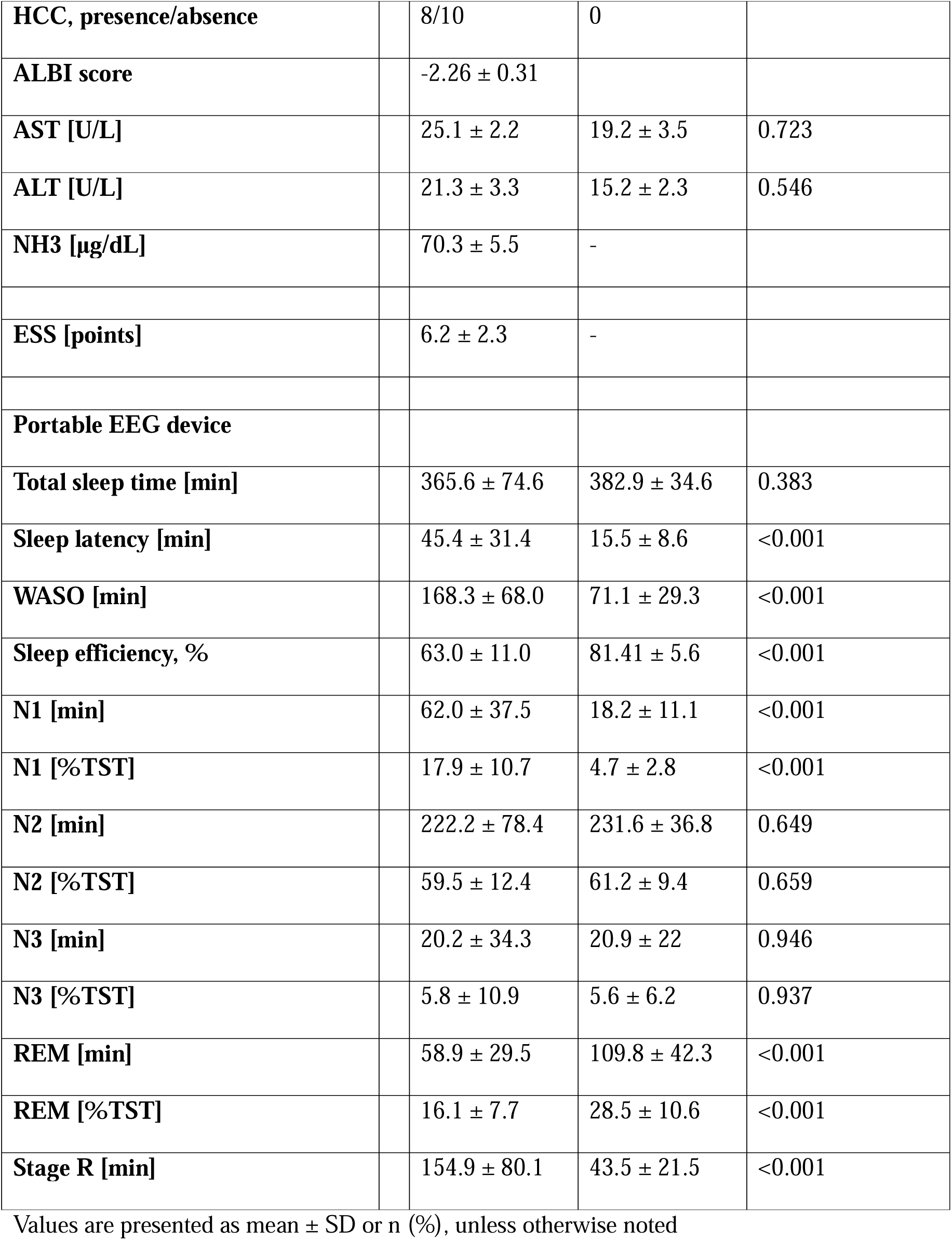
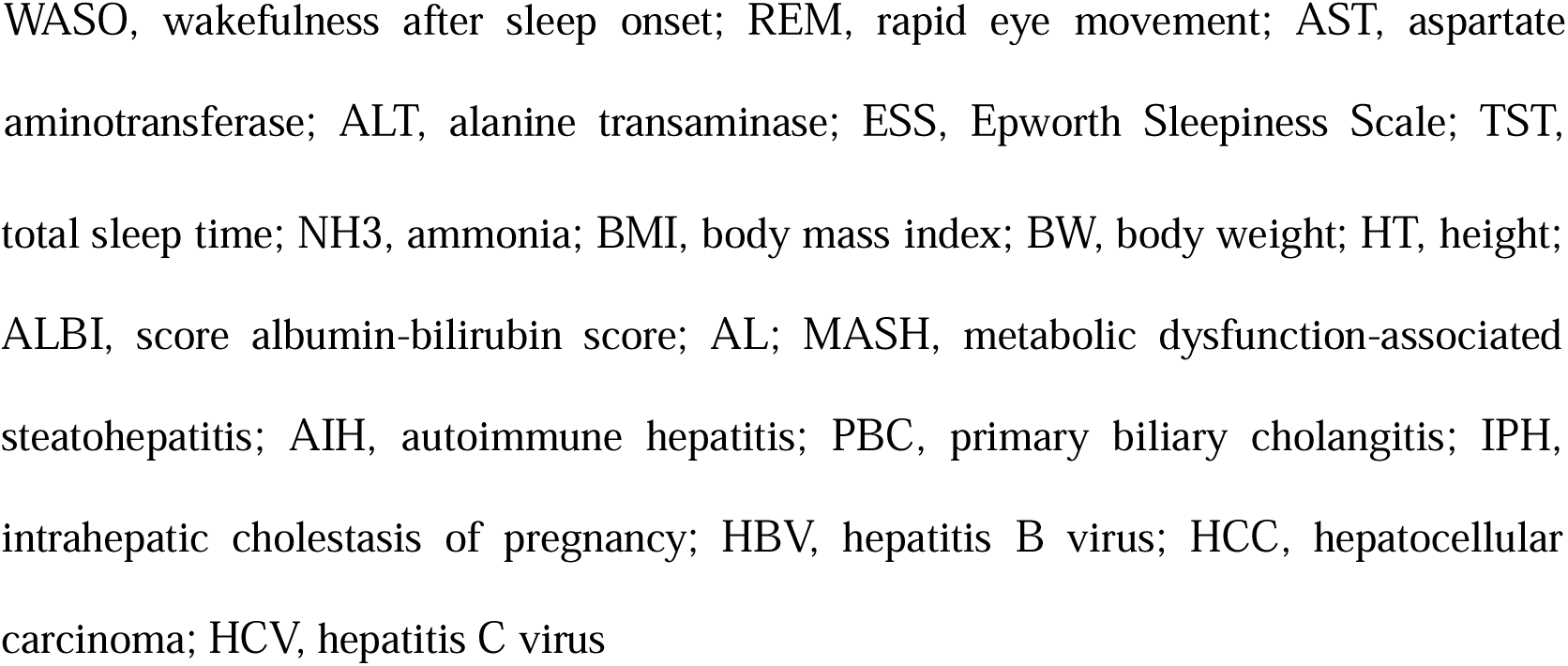
Comparison of Sleep Parameters in Patients with Liver Cirrhosis and Healthy Controls.

|  | **Patients with**<br>**liver cirrhosis**<br>**(n = 18)** | **Healthy**<br>**controls**<br>**(n = 18)** | **P-values** |
| --- | --- | --- | --- |
| **Age [years]** | 72.9 ± 8.3 | 75.9 ± 3.9 | 0.924 |
| **Sex, male/female** | 15(83%) | 15(83%) | 1 |
| **BMI [kg/m<sup>2</sup>]** | 24.5 ± 4.3 | 23.6 ± 3.3 | 0.945 |
| **Etiology,**<br>**HBV/HCV/MASH/AIH/PBC** | 2/2/12/1/1 |  |  |
| **HCC, presence/absence** | 8/10 | 0 |  |
| **ALBI score** | -2.26 ± 0.31 |  |  |
| **AST [U/L]** | 25.1 ± 2.2 | 19.2 ± 3.5 | 0.723 |
| **ALT [U/L]** | 21.3 ± 3.3 | 15.2 ± 2.3 | 0.546 |
| **NH3 [µg/dL]** | 70.3 ± 5.5 | - |  |
| **ESS [points]** | 6.2 ± 2.3 | - |  |
| **Portable EEG device** |  |  |  |
| **Total sleep time [min]** | 365.6 ± 74.6 | 382.9 ± 34.6 | 0.383 |
| **Sleep latency [min]** | 45.4 ± 31.4 | 15.5 ± 8.6 | <0.001 |
| **WASO [min]** | 168.3 ± 68.0 | 71.1 ± 29.3 | <0.001 |
| **Sleep efficiency, %** | 63.0 ± 11.0 | 81.41 ± 5.6 | <0.001 |
| **N1 [min]** | 62.0 ± 37.5 | 18.2 ± 11.1 | <0.001 |
| **N1 [%TST]** | 17.9 ± 10.7 | 4.7 ± 2.8 | <0.001 |
| **N2 [min]** | 222.2 ± 78.4 | 231.6 ± 36.8 | 0.649 |
| **N2 [%TST]** | 59.5 ± 12.4 | 61.2 ± 9.4 | 0.659 |
| **N3 [min]** | 20.2 ± 34.3 | 20.9 ± 22 | 0.946 |
| **N3 [%TST]** | 5.8 ± 10.9 | 5.6 ± 6.2 | 0.937 |
| **REM [min]** | 58.9 ± 29.5 | 109.8 ± 42.3 | <0.001 |
| **REM [%TST]** | 16.1 ± 7.7 | 28.5 ± 10.6 | <0.001 |
| **Stage R [min]** | 154.9 ± 80.1 | 43.5 ± 21.5 | <0.001 |

Values are presented as mean ± SD or n (%), unless otherwise noted
WASO, wakefulness after sleep onset; REM, rapid eye movement; AST, aspartate
aminotransferase; ALT, alanine transaminase; ESS, Epworth Sleepiness Scale; TST,
total sleep time; NH3, ammonia; BMI, body mass index; BW, body weight; HT, height; ALBI, score albumin-bilirubin score; AL; MASH, metabolic dysfunction-associated steatohepatitis; AIH, autoimmune hepatitis; PBC, primary biliary cholangitis; IPH, intrahepatic cholestasis of pregnancy; HBV, hepatitis B virus; HCC, hepatocellular carcinoma; HCV, hepatitis C virus

### Sleep quality evaluation

The average data of 3 days were collected for all (100%) patients; If data collection was inadequate, it was repeated on a different day. A feature of this device allows data analysis to be performed immediately upon recharging with a smartphone application. Data were collected from all 20 patients.

No significant differences were observed between the healthy controls and patients with cirrhosis in age, sex, and body mass index. However, patients with cirrhosis had lower albumin-bilirubin scores and higher ammonia levels than the control group.

Fig 3a shows a real good quality case, deep Sleep EEG Score of 910/1000 (placing in the top 5%), with a total sleep duration of 7 hours and 1 minute. Overall sleep quality was 88/100, took 14 minutes to fall asleep, spent 51 minutes awake during the night, and sleep efficiency was 87%. (Fig 3a).

**Fig 3.**
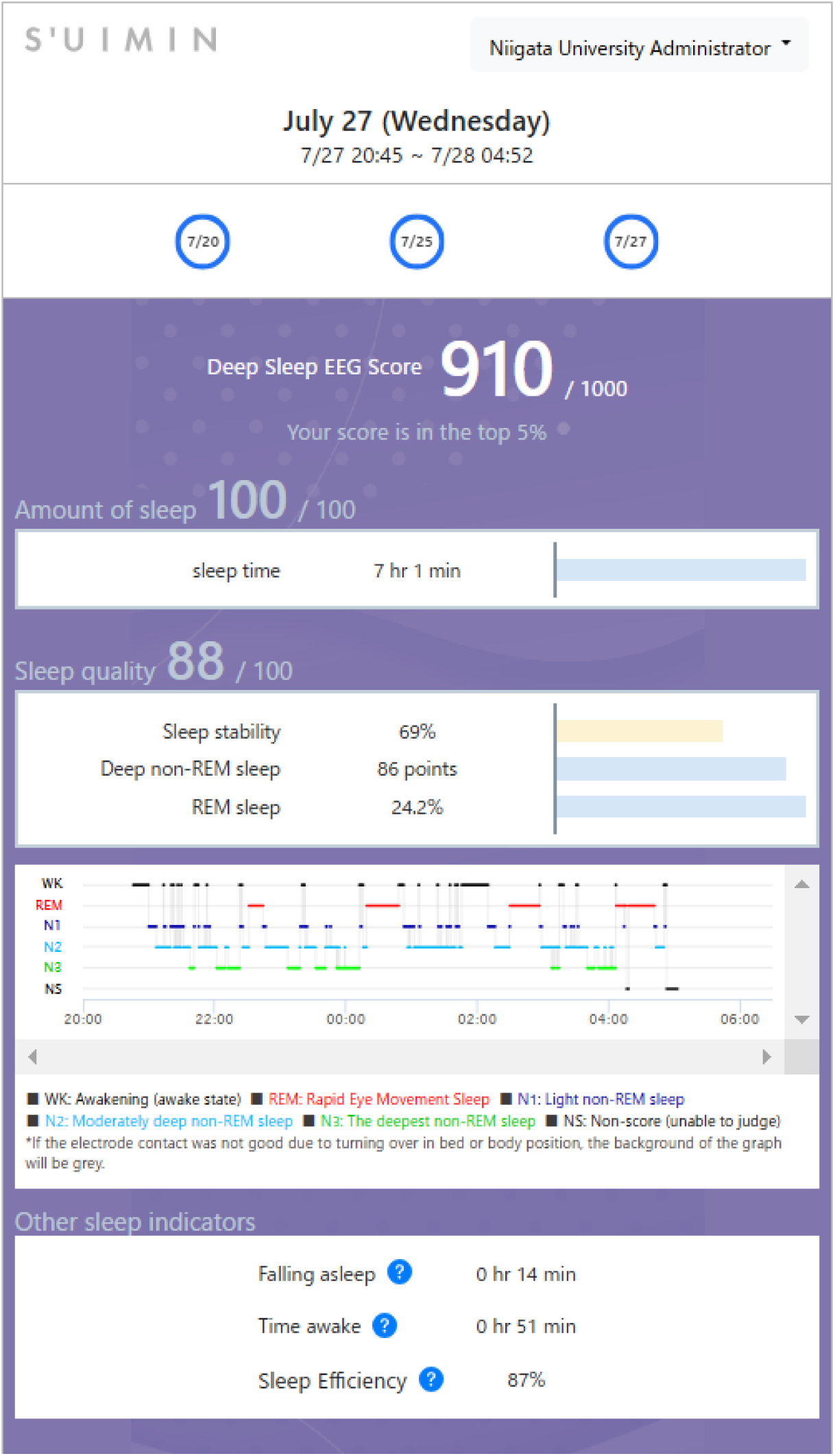
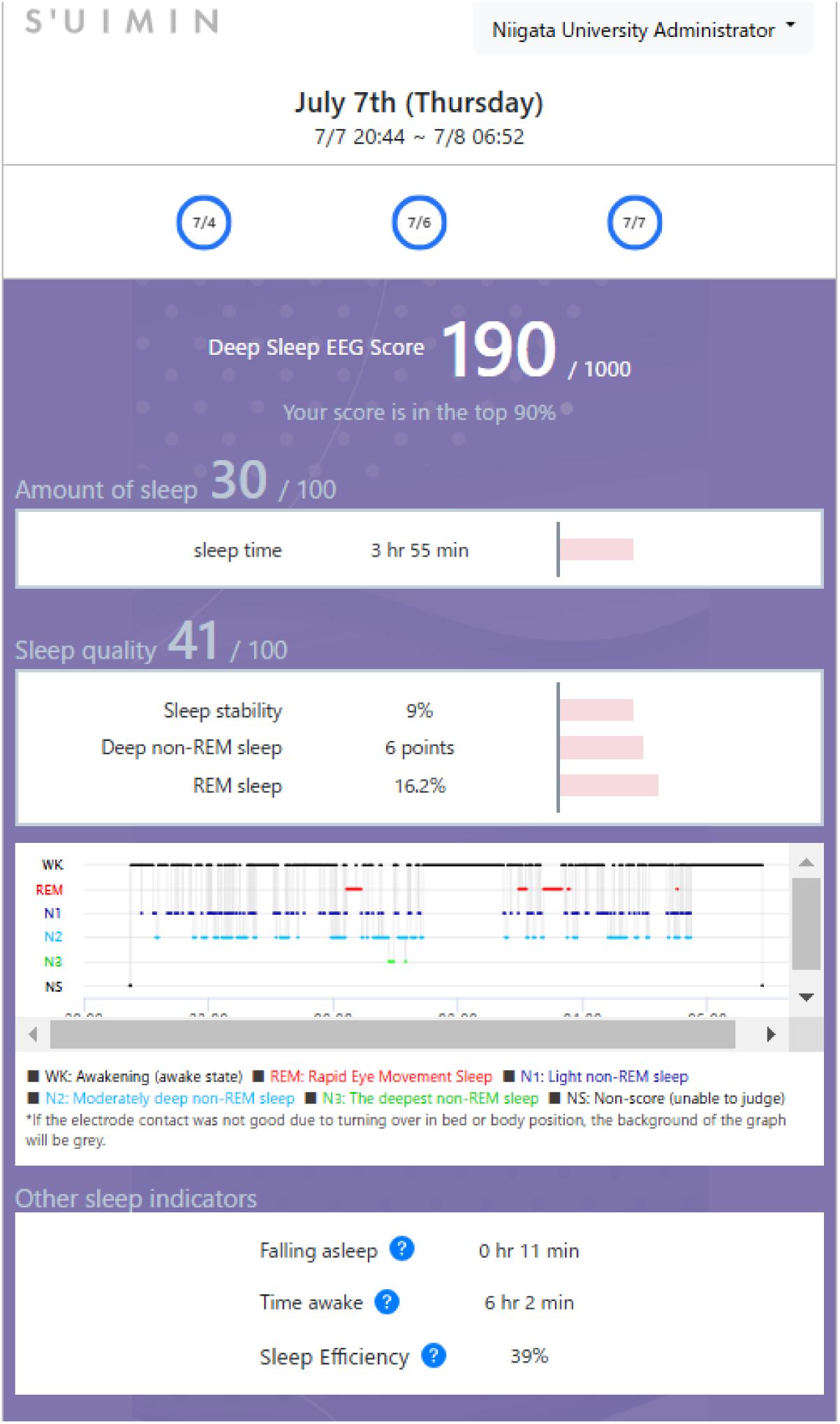
A Smartphone Screen Displaying a.

Fig 3b shows a bad quality case, EEG Score was 190/1000 (placing in the in the bottom 10%), with a total sleep duration of 3 hours and 55 minutes. Overall sleep quality was 41/100 took 11 minutes to fall asleep spent 6 hours and 2 minutes awake during the night, and your sleep efficiency was 39%. (Fig 3b). More than half of the patients experienced a phenomenon known as sleep misrecognition, where they were objectively sleep deprived even though they thought they were actually sleeping well. In the control group, no participant had >11 points on the ESS. However, the portable EEG device revealed a high degree of dissociation between the two groups regarding sleep efficiency. Although 90% of the respondents had scores of ≤5, indicating almost no sleep disturbance, more than half had a decrease in sleep efficiency (Table 3).

### Comparison of sleep variables

Cumulative displays show sleep architecture, illustrating the percentage of individuals in each sleep stage, N1, N2, N3, and REM (Y-axis) according to the time from sleep onset (X-axis) by EEG-based sleep clusters. In patients with cirrhosis, we observed differences in sleep architecture, including the low percentage of N3 in the middle and worse sleep groups and less distinct REM sleep cycles in the worse sleep group (Fig 4).

**Fig 4.**
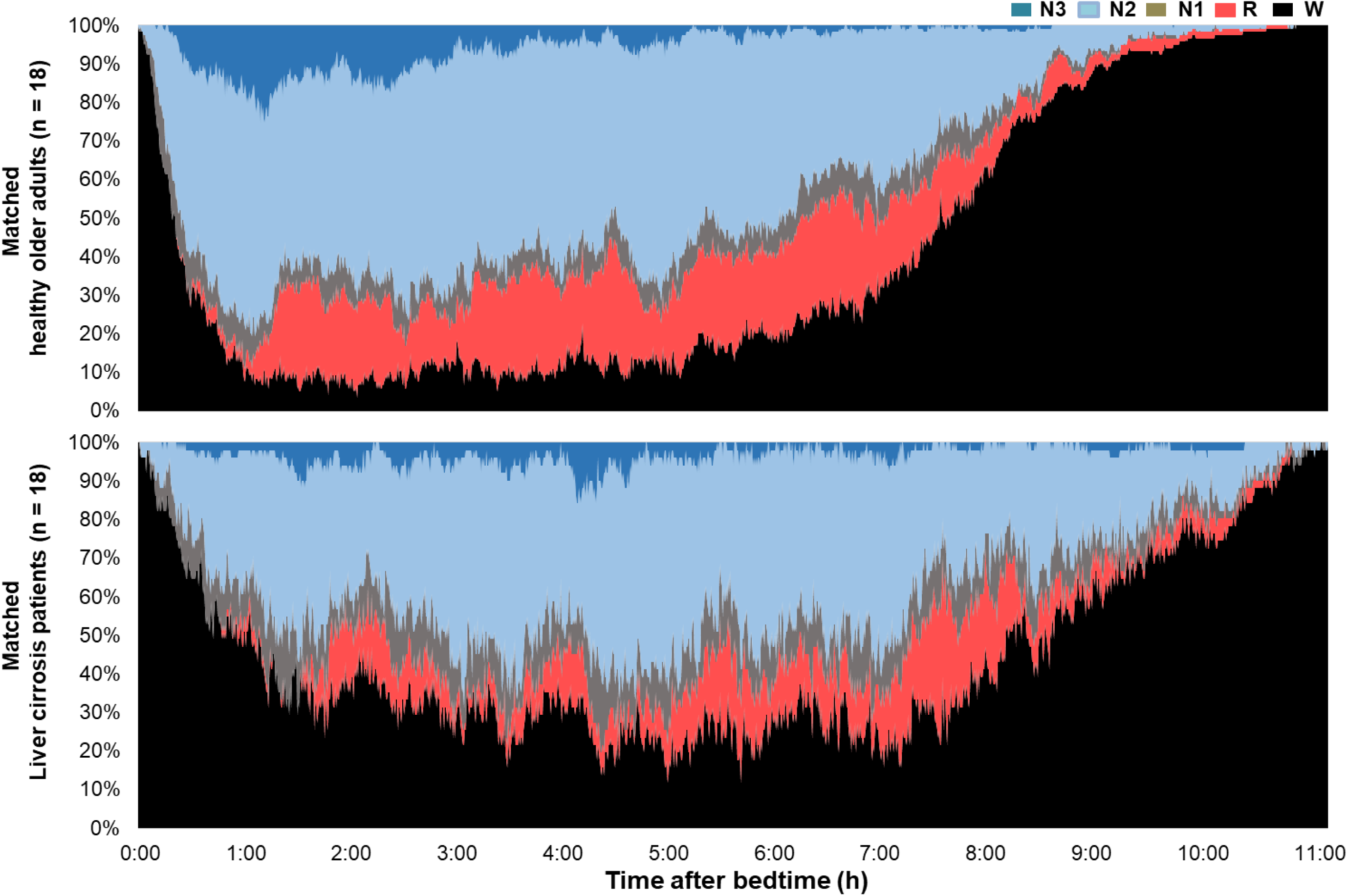
Comparison of Sleep Variables.

Patients with cirrhosis also had a significantly longer sleep latency (45.4 min) and wakefulness after sleep onset (168.3 min) than did healthy controls (15.5 min and 71.1 min, respectively; *P*<0.001), as shown in Table 1 and Fig 1. However, the total sleep time was similar between the groups (356.6 min and 382.9 min, respectively), as presented in Table 1, Fig 4, and Fig 5.

**Fig 5.**
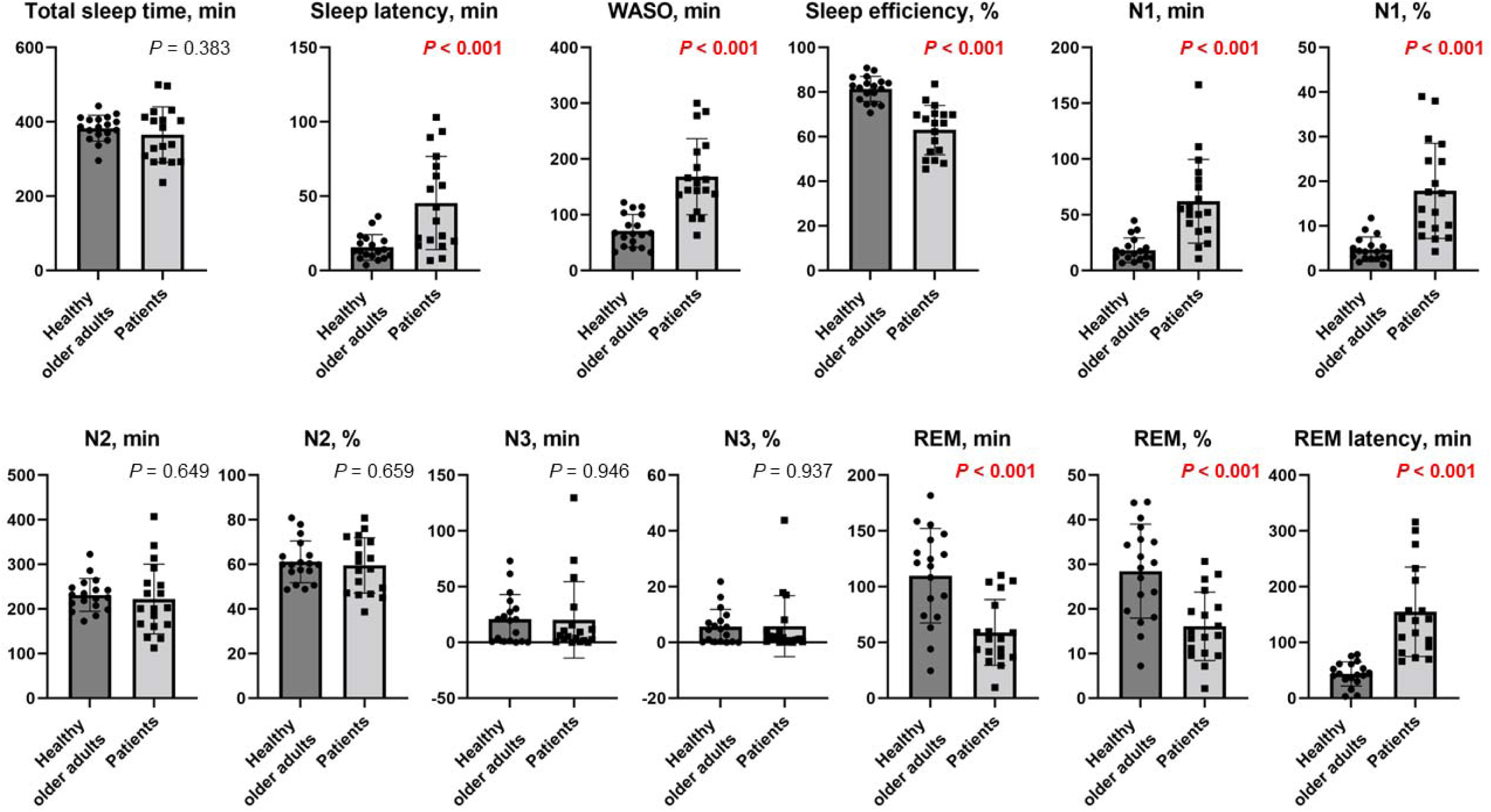
Box and Whisker Diagram Comparing Patients with Cirrhosis at Each Sleep Stage with Healthy Controls.

Regarding sleep structure, compared with healthy controls, patients with cirrhosis had more total time and proportion of awakenings and N1 (62 min), and less total time and proportion of REM (16.1 min), indicating more shallow sleep throughout the night. Additionally, patients with cirrhosis had a markedly long REM latency and decreased total REM time and percentage compared with the healthy controls (*P*<0.001) (Table 3, Fig 5).

## Discussion

This study compared sleep disturbances in patients with early cirrhosis with those in healthy individuals, with exclusion criteria including previously reported causes of sleep disturbances in cirrhosis patients, such as severe hepatic encephalopathy (HE) and itching. Patients with cirrhosis showed longer sleep latency, more frequent awakenings during the night, and increased amounts of N1 sleep than did healthy individuals, exhibiting decreased REM sleep, longer REM latency, and reduced sleep efficiency. Although no significant difference in the amount of N3 sleep was observed between the two groups (Table 3), regarding the temporal changes in its occurrence (Fig 5), patients with cirrhosis showed an absence of N3 sleep during the first 1–2 h after sleep onset compared with healthy adults. Instead, a higher percentage of N1 sleep and wakefulness was observed (Fig 5). According to the non-REM-REM sleep cycle theory, disruption in non-REM sleep by wakefulness resets the cycle before reaching REM sleep, resulting in delayed REM latency and poorer sleep quality (Table 1 and Fig 1). In this study, none of the patients with cirrhosis were identified as having sleep apnea syndrome based on ESS questionnaire assessment. Possible reasons for the observed sleep disturbances include changes in intestinal bacteria, worsening of liver reserve function, chronic inflammation, and psychological burden caused by disease distress, all of which have been reported to contribute to HE, delayed peak secretion, and poor sleep quality. The prevalence of sleep disturbance in patients with cirrhosis ranges between 48% and 55% [14,15]. Sleep-wake cycle reversal is considered an early sign of HE and associated with daytime dysfunction [16]. In animal models and human studies, HE is associated with circadian cycle disturbances. Cirrhosis patients exhibit higher daytime melatonin levels and lower nighttime melatonin clearance, as altered melatonin secretion patterns are associated with delayed peak secretion and sleep quality [17]. Changes in intestinal bacteria, worsening with each reserve, chronic inflammation, and psychological burden due to disease distress could also cause HE and delayed peak secretion and affect sleep quality [18]. Furthermore, primary biliary cholangitis, itching, sleep disturbances, and chronic liver diseases, including hepatitis and cirrhosis, can cause itching as symptoms progress. Intense itching can make sleeping at night difficult. Approximately 70% of patients with primary biliary cholangitis experience sleep disturbances [8]. To date, the ESS has provided an unequivocal survey tool [19], as studies have consistently used it as a subjective instrument to assess daytime sleepiness in patients with chronic liver disease, with comparable results [20,21]. Many of these studies were published in Athens, and most were based on the quantification of sleep disturbances using sleep questionnaires. In previous research, sleep progressed from non-REM sleep stages 1, 2, and 3 to REM sleep, with each cycle lasting approximately 90 minutes and being repeated 3–5 times a night, depending on the individual [22]. In the first half of sleep, non-REM sleep 3 was more frequent, and REM sleep increased in the second half. These sleep categories were analyzed based on brain waves, with amplitude, including fast and slow waves, and alpha, beta, and delta waves used to define shallow or deep sleep. With aging, sleep changes occur, including decreased deep sleep (non-REM sleep 3) and increased percentage of shallow sleep [23]. Thus, in a study population of older individuals, adjusting for age, sex, race, and environment is necessary when comparing them with patients with cirrhosis. This study examined poor sleep efficiency in patients with cirrhosis compared with healthy controls by analyzing the same model in Japanese participants of the same age, sex, and body size while using the same equipment. Despite the high prevalence of sleep disorders in patients with cirrhosis, evidence regarding their management remains limited. HE and sleep disorders may have overlapping mechanisms of action. Therefore, in a patient with both cirrhosis and daytime sleepiness with hyperammonemia, treating hyperammonemia first is the most reasonable action [16,24]. Furthermore, melatonin metabolism is impaired in patients with cirrhosis; therefore, melatonin administration could improve their sleep quality and decrease daytime sleepiness [25]. Behavioral therapies, including exposure to bright light early in the morning and avoidance of bright light at night, may also aid treatment [26]. However, data and results are limited, as this study included a small sample. Therefore, the treatment of sleep disturbances in patients with cirrhosis should be further investigated. Sleep characteristics should be monitored, and adequate treatment should be provided to patients with sleep disturbances.

### Limitations

This study has some limitations. First, the small sample size may limit the generalizability of the findings, emphasizing the need for larger studies. Although this study provides valuable insights, unlike previous studies that relied primarily on questionnaires, this study utilized innovative scientific and objective devices to reveal latent sleep disorders in healthy individuals and those in the early stages of liver cirrhosis. Based on these findings, further large-scale investigations are anticipated in the future.

## Conclusion

This study sheds light on the significant sleep disturbances experienced by patients with early-stage cirrhosis, compared with healthy controls, and highlights the importance for addressing these disturbances as part of cirrhosis management. By employing advanced portable EEG technology and adhering to standardized sleep architecture scoring criteria, the research provides a comprehensive analysis of sleep patterns, revealing key differences such as increased sleep latency, frequent nighttime awakenings, reduced REM sleep, and a higher proportion of shallow sleep (N1) among patients with cirrhosis. These findings emphasize the prevalence of sleep misrecognition, wherein patients with cirrhosis perceive their sleep as adequate despite objectively poor sleep efficiency.

The paper also identified potential contributors to these sleep disruptions, including hyperammonemia, altered melatonin metabolism, and disease-related psychological burdens, all of which may have overlapping mechanisms with HE. Although highlighting the clinical relevance of these findings, this research underscores the need for multifaceted management strategies, including treating hyperammonemia, administering melatonin, and adopting behavioral therapies such as light exposure modifications, to improve sleep quality and daytime functionality in patients with cirrhosis.

## Data Availability

All data produced in the present study are available upon reasonable request to the authors

## Abbreviations

AASM: American Academy of Sleep Medicine
EEG: Electroencephalogram
ESS: Epworth Sleepiness Scale
HE: Hepatic encephalopathy
non-REM: non-rapid eye movement
REM: rapid eye movement

## Acknowledgments

The authors thank S’UIMIN Inc. for excellent assistance with analysis by sleep technologists and machine analysis.

## Notes

### Competing Interest Statement

The authors have declared no competing interest.

### Funding Statement

grant-in-aid for scientific research (grant number: 20K08326) from the Ministry of Education, Culture, Sports, Science, and Technology (given to Hiroteru Kamimura).

